# Quantification of different iron forms in the aceruloplasminemia brain to explore iron-related neurodegeneration

**DOI:** 10.1101/2020.10.15.20206102

**Authors:** Lena H.P. Vroegindeweij, Lucia Bossoni, Agnita J.W. Boon, J.H. Paul Wilson, Marjolein Bulk, Martina Huber, Jacqueline Labra-Muñoz, Andrew Webb, Louise van der Weerd, Janneke G. Langendonk

**Author notes:** Both authors contributed equally to the manuscript (Lena H.P. Vroegindeweij, Lucia Bossoni). **Corresponding author:** Lucia Bossoni, PhD, Leiden University Medical Center, PO Box 9600, 2300 RC Leiden, The Netherlands, | E.

## Abstract

**Introduction:** Aceruloplasminemia is an ultra-rare neurodegenerative disorder associated with massive brain iron accumulation. It is unknown which molecular forms of iron accumulate in the brain of patients with aceruloplasminemia. As the disease is associated with at least a fivefold increase in brain iron concentration compared to the healthy brain, it offers a unique model to study the role of iron in neurodegeneration and the molecular basis of iron-sensitive MRI contrast.

**Methods:** The iron-sensitive MRI metrics inhomogeneous transverse relaxation rate (R_2_*) and magnetic susceptibility obtained at 7T were combined with Electron Paramagnetic Resonance (EPR) and Superconducting Quantum Interference Device (SQUID) magnetometry to specify and quantify the different iron forms per gram wet-weight in a post-mortem aceruloplasminemia brain, with focus on the basal ganglia, thalamus, red nucleus, dentate nucleus, superior-and middle temporal gyrus and white matter. MRI, EPR and SQUID results that had been previously obtained from the temporal cortex of healthy controls were included for comparison.

**Results:** The brain iron pool in aceruloplasminemia consisted of EPR-detectable Fe^3+^ ions, magnetic Fe^3+^ embedded in the core of ferritin and hemosiderin (ferrihydrite-iron), and magnetic Fe^3+^ embedded in oxidized magnetite/maghemite minerals (maghemite-iron). Of all the studied iron pools, above 90% was made of ferrihydrite-iron, of which concentrations up to 1065 µg/g were detected in the red nucleus. Although deep gray matter structures in the aceruloplasminemia brain were three times richer in ferrihydrite-iron than the temporal cortex, ferrihydrite-iron in the temporal cortex of the patient with aceruloplasminemia was already six times more abundant compared to the healthy situation (162 µg/g vs. 27 µg/g). The concentration of Fe^3+^ ions and maghemite-iron were 1.7 times higher in the temporal cortex in aceruloplasminemia than in the control subjects. Of the two quantitative MRI metrics, R_2_* was the most illustrative of the pattern of iron accumulation and returned relaxation rates up to 0.49 ms^-1^, which were primarily driven by the abundance of ferrihydrite-iron. Maghemite-iron did not follow the spatial distribution of ferrihydrite-iron and did not significantly contribute to MRI contrast in most of the studied regions.

**Conclusions:** Even in extremely iron-loaded cases, iron-related neurodegeneration remains primarily associated with an increase in ferrihydrite-iron, with ferrihydrite-iron being the major determinant of iron-sensitive MRI contrast.

## Introduction

Aceruloplasminemia (OMIM #604290) is a severe adult-onset form of Neurodegeneration with Brain Iron Accumulation (NBIA), caused by homozygous or compound heterozygous mutations in the ceruloplasmin (*CP*) gene. Iron accumulation in aceruloplasminemia has been directly linked to its genetic background, as ceruloplasmin-mediated oxidation of ferrous iron (Fe^2+^) to ferric iron (Fe^3+^) is a prerequisite for transport of iron across cell membranes and for binding to transferrin (1). As a result of ceruloplasmin dysfunction in the brain leading to insufficient cellular iron efflux, iron accumulates within perivascular astrocytes, with total iron concentrations reaching over 600 µg/g in the basal ganglia (2), which is at least five times the normal concentration (3, 4). However, the exact role of different iron forms in the pathophysiology and clinical phenotype of aceruloplasminemia is still not clear. Molecular specification of the massive iron pool in the aceruloplasminemia brain can aid our understanding of iron related neurodegeneration, and perhaps improve therapeutic considerations, which are currently based on iron chelation.

Among the molecular forms in which iron can be found in the brain, three are particularly relevant. In the healthy human brain, iron is predominantly stored within the core of ferritin and hemosiderin, in the form of Fe^3+^-containing ferrihydrite nanocrystals (Fe_2_O_3_•0.5H_2_O) (5). These iron stores are generally regarded as non-damaging and are required for normal brain functions. In contrast, Fe^2+^ ions taking part in the labile iron pool have been associated with iron-related neuronal damage through generation of free radicals via the Fenton reaction (1, 5, 6). The iron-oxide magnetite (Fe_3_O_4_) is a carrier of Fe^2+^ and might thus play a prominent role in iron-related neurodegeneration (5, 7). Histochemical studies in aceruloplasminemia have indicated an abundance of ferritin-bound iron (8, 9), and smaller amounts of Fe^2+^ carriers (8, 10). However, no quantitative studies have been performed to further determine the molecular forms of iron and their distribution throughout the aceruloplasminemia brain.

Iron-sensitive MRI techniques have been commonly applied as an indirect measure of brain iron content in neurodegenerative disorders, using the effect that (para)magnetic iron has on the proton nuclear relaxation rate (R_2_*) and on the magnetic susceptibility of the tissue, which can be measured by Quantitative Susceptibility Mapping (QSM) (11, 12). Although R_2_* mapping is recognized as a robust method to infer iron accumulation in brain tissue, it is affected by several confounders such as cell water content, the macromolecular pool and myelin. In particular, myelin and iron have the same effect on R_2_* values (13). QSM, on the other hand, has the potential to differentiate between myelin and iron, based on their opposite effects on tissue susceptibility (14), and seems less affected by the cellular localization of iron (15). Both iron-sensitive MRI techniques have been used for the quantitative assessment of iron in aceruloplasminemia brain tissue (16, 17), but only provided total iron concentrations and not their specification (18).

The primary aim of this study was to directly quantify different molecular iron forms in the aceruloplasminemia brain. For this purpose, we performed Electron Paramagnetic Resonance (EPR) and Superconducting Quantum Interference Device (SQUID) magnetometry on ten brain regions derived from a post-mortem aceruloplasminemia brain. It should be stressed that, due to the rarity of the disease, aceruloplasminemia brain material is extremely scarce. We present a quantitative overview of Fe^3+^ ions detected by EPR, and magnetic Fe^3+^ embedded in ferritin cores or hemosiderin(ferrihydrite-iron) and oxidized magnetite/maghemite minerals (maghemite-iron), as detected by magnetometry. R_2_* and QSM maps from the same regions are shown with the additional aim to illustrate the extent to which different iron forms affect iron-sensitive MRI metrics. Since MRI is the major imaging modality capable of studying brain iron accumulation *in vivo*, further specification of the molecular basis of iron-sensitive MRI contrast is key to gain more in-depth pathophysiological insights into different clinical stages of iron-related neurodegeneration.

## Methods

### Brain tissue selection

Formalin-fixed brain slices of one end-stage aceruloplasminemia patient (male, age range: 50-55) (19-22) were obtained from the Department of Pathology, Erasmus MC, University Medical Center Rotterdam, The Netherlands. This study has received IRB approval by the Erasmus MC Medical Ethical Committee (MEC-2011-525). The brain slices had been stored in formalin for 5 years. Tissue blocks of approximately 30 x 30 x 10 mm^3^ were dissected, including the basal ganglia, thalamus, red nucleus, dentate nucleus, superior- and middle temporal gyrus and white matter. Ceramic tools were used to prevent metal contamination of the samples.

### MRI data acquisition and data analysis

Before MRI, the dissected tissue blocks were rehydrated in phosphate buffered saline (PBS) for 24 hours and immersed in a proton-free solution (Fomblin LC08, Solvay). ICP-MS detected iron traces up to 3.5 µg/ml in the formalin solution, likely due to leakage of iron from the tissue over time.

MRI scans were performed at room temperature on a 7 T preclinical scanner (Bruker Biospin, Ettlingen, Germany) using a 38-mm quadrature RF coil. For each tissue sample a three-dimensional multi-gradient-echo sequence (MGE) was used with eight equally spaced echoes. First echo-time was 1.96 ms, echo spacing was 2.21 ms, repetition time was 130.4 ms, flip angle was 25º, 150 µm^3^ isotropic spatial resolution. 20 averages were acquired. The total acquisition time was 3h 54minutes.

R_2_* (1/T_2_*) maps were obtained by fitting the signal decay of the magnitude images to a mono-exponential decay function. Susceptibility values (χ) were obtained from the eight-echoes phase images of the MGE data, with the STI-Suite software (version 2.2)(23). Phase unwrapping of the measured phase images and removal of the background field was done with the iHARPERELLA algorithm (24), while magnetic susceptibility estimation and streaking artifact correction was performed with the ‘iLSQR’ algorithm (25). Raw susceptibility values are reported, as no reference region was available in these small tissue samples. Regions of interest (ROIs) were manually drawn within each deep gray matter region, middle temporal gyrus and temporal white matter. For each ROI, mean R_2_* and magnetic susceptibility values were reported.

Additional MR images that had been previously acquired (26) on tissue blocks containing the temporal cortex and the striatum of two control subjects are used as references to illustrate the effect of iron accumulation in the aceruloplasminimia brain on MRI maps. These control tissue blocks had been scanned with an acquisition protocol as described above, with the exception of the echo timing, which was increased as the iron concentration was much lower in these control cases: the first echo time was 12.5 ms long, and the inter-echo time was 10.7 ms.

### EPR experiments and spectra fitting

After MRI, subsections of approximately 40 mg were taken from each gray and white matter structure in the tissue blocks (Figure 1). The resulting subsections were prepared for EPR as previously described (27). The 9 GHz continuous wave EPR measurements were performed at 6 K using an ELEXSYS E680 spectrometer (Bruker, Rheinstetten, Germany) equipped with a rectangular cavity. The microwave frequency was 9.4859 GHz, modulation frequency 90 kHz, power attenuation 20 dB, receiver gain 60 dB and modulation amplitude 6 Gpp. Power attenuation was chosen after a progressive power saturation experiment (Supplementary material, Figure S1). The accumulation time was approximately five minutes per spectrum.

**Figure 1.**
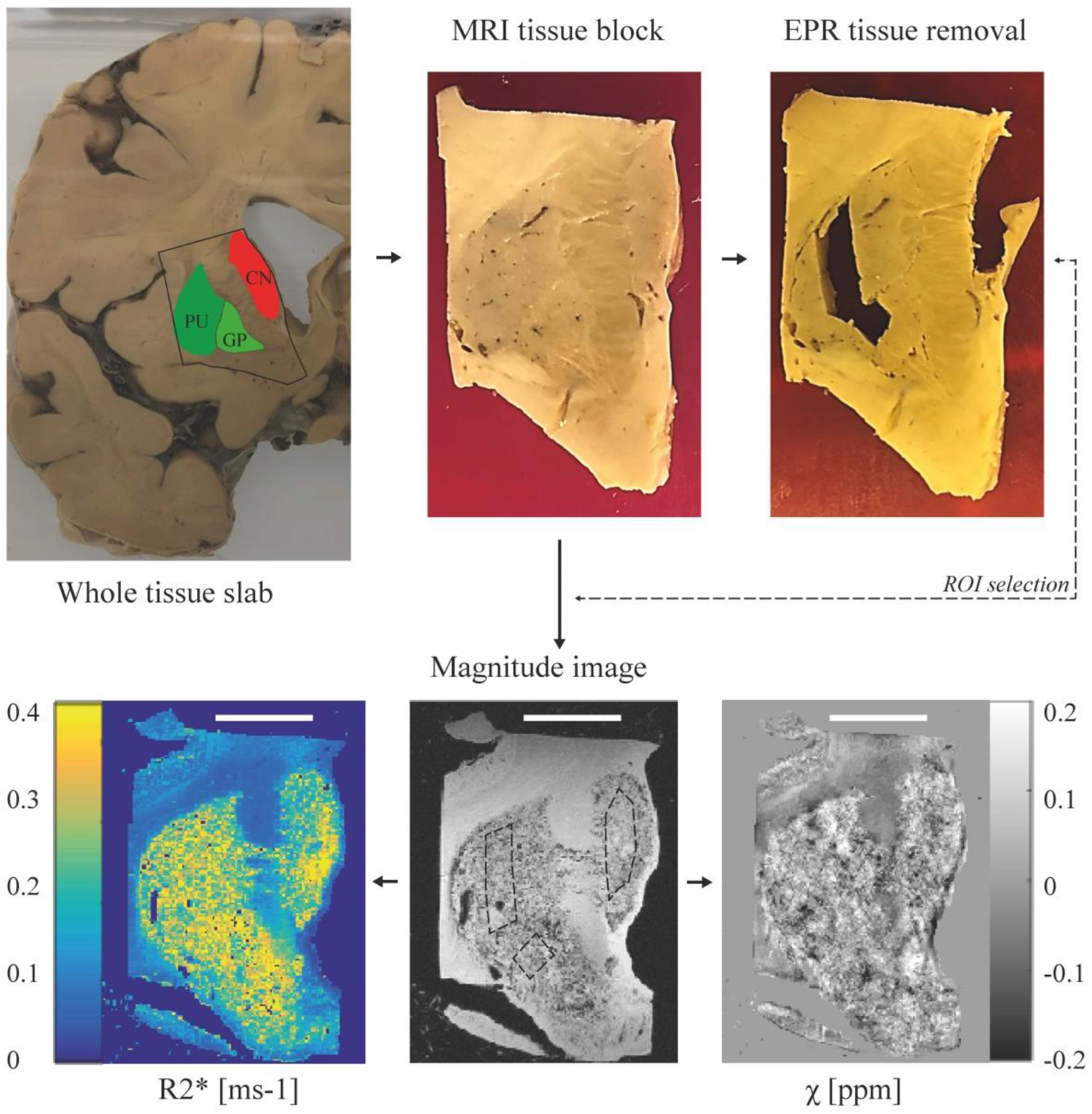
Pipeline of the experimental protocol. In this example, the basal ganglia (caudate nucleus – CN, putamen – PU, globus pallidus – GP), were dissected from the appropriate brain slab and prepared for MRI. Following MRI, small subsections were taken from each basal ganglion structure and prepared for EPR. Regions of interest (ROIs) on the MRI were matched with the EPR sampling locations. For SQUID, subsections of tissue were taken adjacent to the sampling locations for EPR. Scale bar in the bottom figures, 1 cm.

Figure 2 shows a baseline corrected EPR spectrum of the tissue block extracted from the putamen. While the spectra consisted of several bands, in this work, we focused on the low field g’=4.3 band, which originates from high-spin Fe^3+^ complexed in a site of low symmetry (27). A close look at the temperature dependence of the band suggests the presence of a weak antiferromagnetic coupling (Supplementary material, Figure S2).

**Figure 2.**
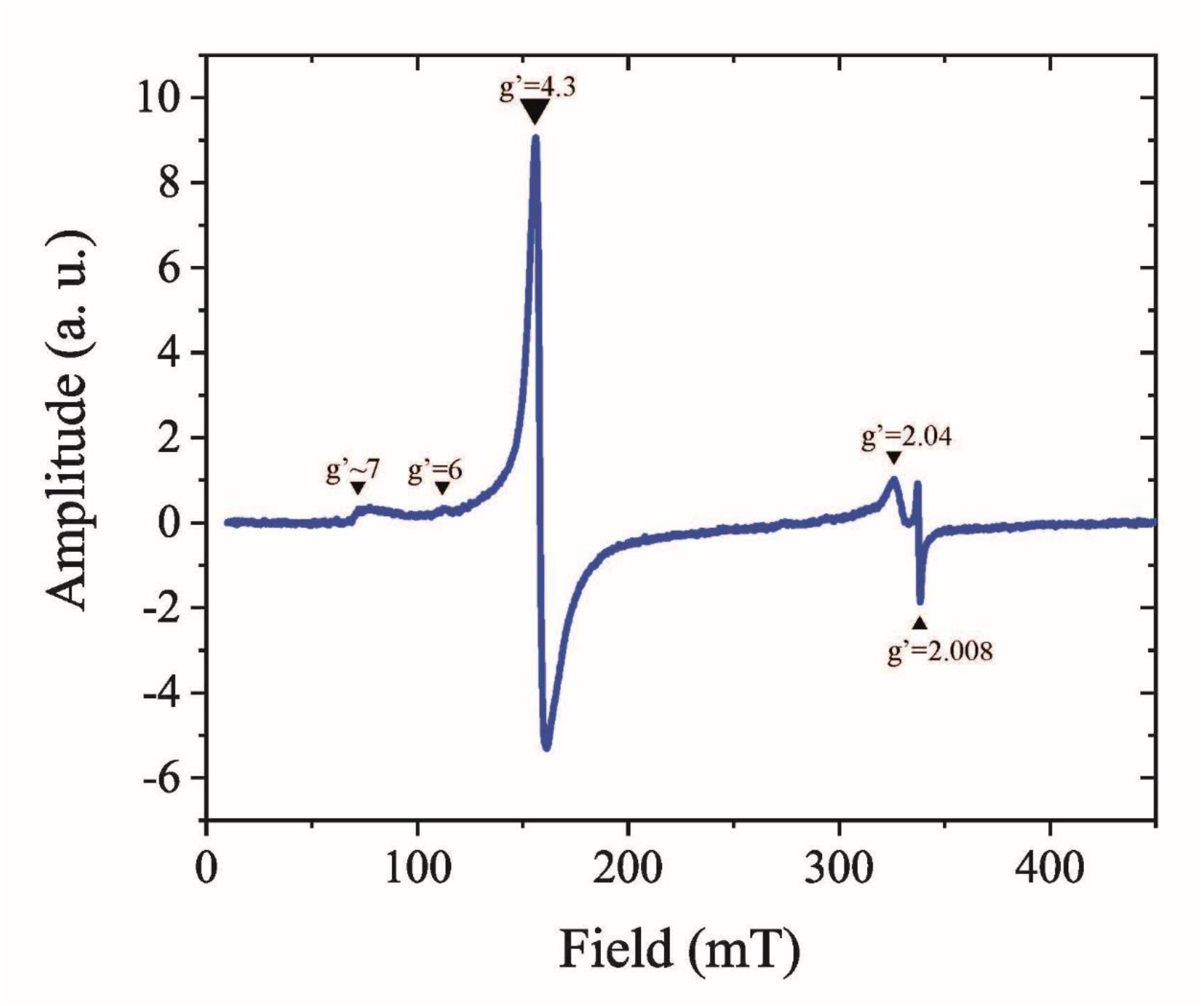
Determination of Fe^3+^ ions by Electron Paramagnetic Resonance (EPR) at 6 K. First derivative, baseline corrected EPR spectrum of the putamen. The spectrum consists of several bands: a g’∼7 poorly defined signal, a weak band at g’=6, an intense band at g’=4.3, a largely asymmetric band around g’=2.04, and a narrow band at g’=2.008.

The g’=4.3 bands detected in each sample were fit to the spin Hamiltonian which describes high-spin Fe^3+^ (27). EPR spectra simulations and fitting were done with EasySpin version 5.2.28 (28). Fitting parameters and examples of the fitted spectra are shown in the Supplementary material (Table S1 and Figure S3). The Fe^3+^ concentration of each sample was calculated by taking the second integral of the fitted spectrum and comparing it to the second integral of a reference solution Fe-EDTA of known Fe^3+^ concentration (35.7 µM).

### Magnetometry experiments

Following EPR experiments, subsections were taken from the caudate nucleus, putamen, substantia nigra, thalamus, red nucleus, dentate nucleus, temporal cortex and white matter, mostly adjacent to the sampling locations for EPR. The total volume of specimens for each brain region was approximately 10 x 10 x 10 mm^3^. The globus pallidus was not included because of its smaller volume. Tissue samples were removed from formalin, frozen in liquid nitrogen, and freeze-dried. The brain material was subsequently pelleted and placed in the sample holder of a Superconducting Quantum Interference Device (SQUID) magnetometer.

Isothermal Remanent Magnetization (IRM) curves were acquired with a Quantum Design MPMS-XL SQUID magnetometer with the Reciprocating Sample Option (RSO) probe, as reported previously (18, 27), to explore whether the samples retained magnetization outside of an external magnetic field following stepwise exposure to higher magnetic field strengths at a constant temperature (29, 30). Two IRM curves were acquired: one at 5 K, which can report on ferrihydrite, the mineral found in the core of ferritin proteins and hemosiderin aggregates; and a second at 100 K, which saturated around 250 mT, indicative of magnetite or its oxidation product, maghemite. An example of the IRM curves for one of the samples is shown in Figure 3. The IRM values were obtained after dividing the raw magnetic moment by the wet weight (ww) of the sample. Saturation IRM values (SIRM) were extracted by fitting the IRM curves to an offset-corrected Langevin model, as in our previous work (18).

**Figure 3.**
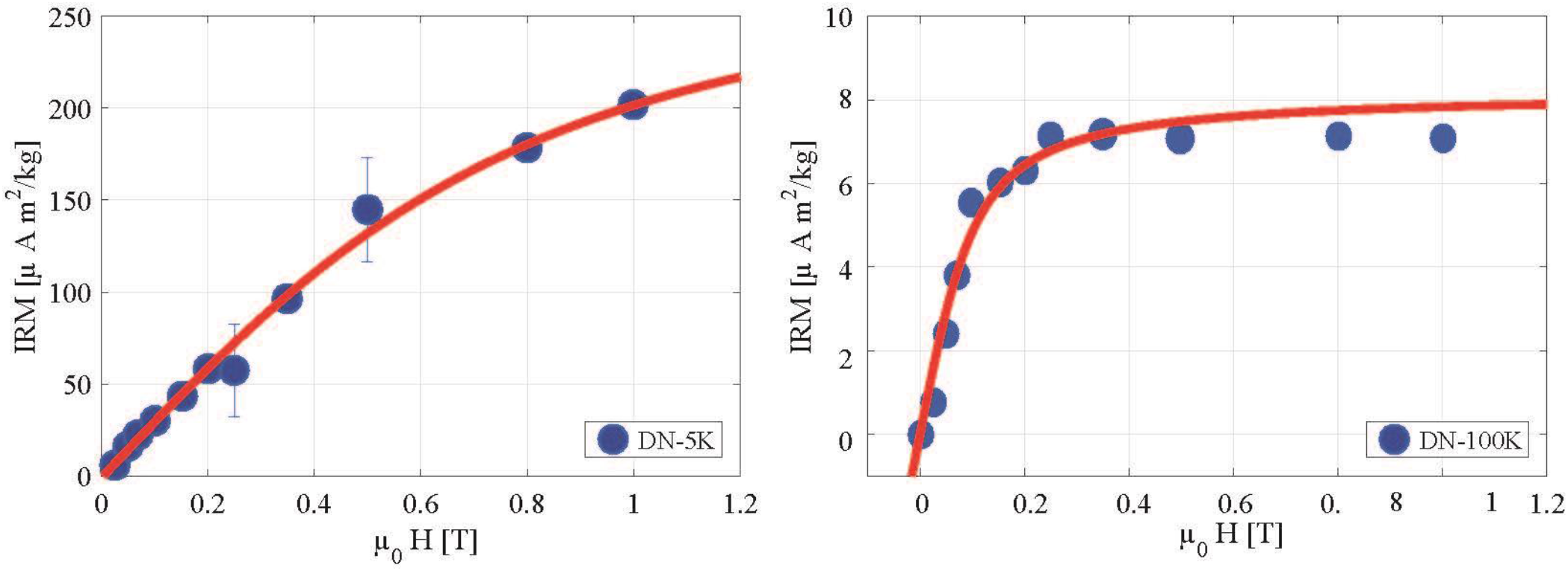
Determination of ferrihydrite and maghemite by Isothermal Remanent Magnetization (IRM). IRM curves of the dentate nucleus (DN) at 5K (left) and 100 K (right). The IRM curve at 100 K saturates around 0.25 T, which is a strong indication for the presence of magnetite or maghemite.

Ferrihydrite and maghemite concentrations were estimated by dividing the SIRM values by the saturation magnetization (M_s_) of the specific mineral: M_s_= 0.62 Am^2^/kg for ferrihydrite (31), and 74 Am^2^/kg for maghemite (32). Maghemite was chosen instead of magnetite, due to the high chance of sample oxidation during tissue processing. Subsequently, the ferrihydrite-iron and maghemite-iron concentrations were calculated by multiplying the mineral concentration by the iron mass-ratio for the given mineral. Concentrations of Fe^3+^, ferrihydrite-iron and maghemite-iron that had been previously obtained under identical conditions from the temporal cortex of healthy controls are provided for comparison (18).

Raw data post-processing of MRI, EPR and SQUID data was performed with an in-house written code in MatlabR2016a. Raw data obtained in this study are downloadable at the link: http://dx.doi.org/10.17632/8b3fvc3h3f.1

## Results

Table 1 provides an overview of all data collected from the aceruloplasminemia brain. The highest R_2_* values were found in the red nucleus, lateral division of the thalamus and substantia nigra. On average, R_2_* values within deep gray matter structures in aceruloplasminemia were over two times higher than those obtained from the temporal cortex. The deep gray matter structures in aceruloplasminemia had at least a three-fold increase in relaxation rates with respect to the close-by white matter, while in the temporal cortex the relative gray matter/white matter relaxation rate was reversed. Higher relaxation rates were observed in the white matter, which indicates a large fraction of iron being embedded in myelin. In the control subjects, both the white matter of the internal capsule and of the temporal lobe showed higher relaxation rates compared to the close-by gray matter (Figure 4), but in all regions the relaxation rates were substantially lower than for the aceruloplasminemia case. Susceptibility values in aceruloplasminemia were only partially in accordance with the pattern of iron accumulation as suggested by the R_2_* maps and were more heterogeneous, within the structures (Figure 4). Notable exceptions were the dentate nucleus, parts of the putamen and the temporal white matter, where a striking paramagnetic component was observed compared to close-by structures.

**Table 1.**
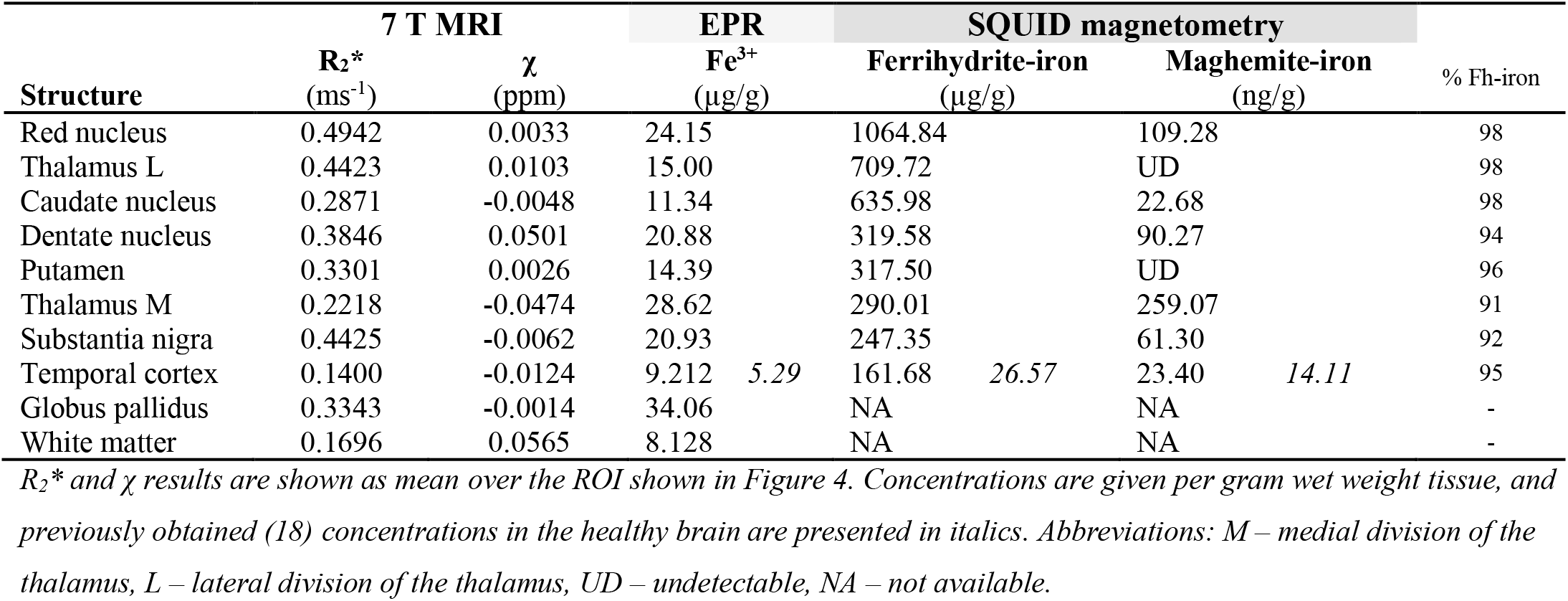
Overview of MRI, EPR and SQUID results obtained from the aceruloplasminemia brain.

**Figure 4.**
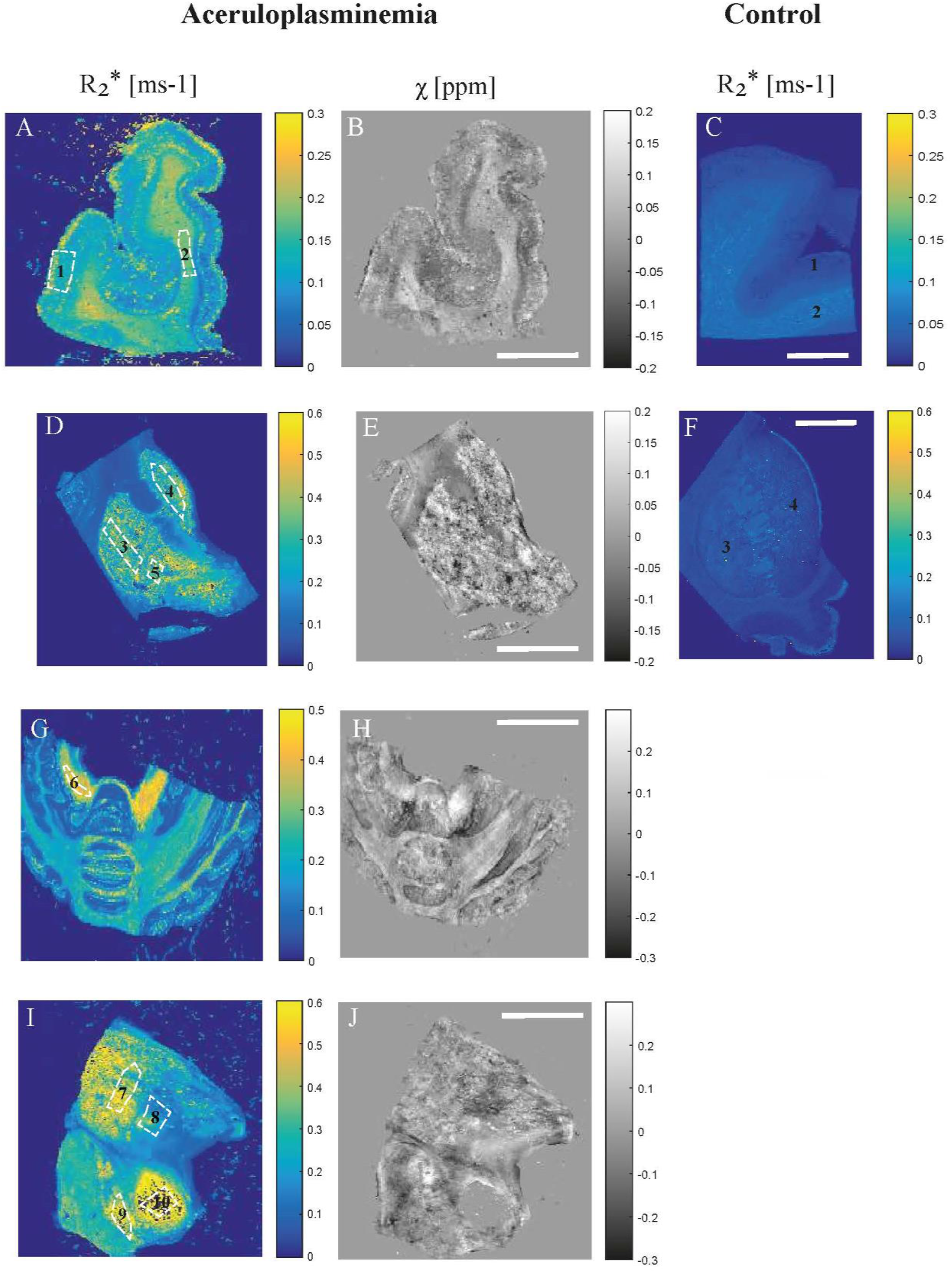
Comparison of quantitative MRI results. The R_2_* and QSM maps are shown with the outlined regions of interest (ROIs) in the temporal cortex (1) and white matter (2) (A, B), putamen (3), caudate nucleus (4) and globus pallidus (5) (D, E), dentate nucleus (6) (G, H), thalamus (7 – lateral division, 8 – medial division), substantia nigra (9) and red nucleus (10) (I, J) in aceruloplasminemia. Note that for the red nucleus, in which the highest R_2_* values were found, the susceptibility signal was predominantly masked out (J). R_2_* maps of the temporal cortex (1) and white matter (2) (C), putamen (3) and caudate nucleus (4)(F) of a control subject are provided for comparison. Scalebar, 1 cm.

Ferrihydrite-iron was the most abundant form of iron in all investigated brain regions in aceruloplasminemia and represented 91-98% of all iron (Table 1). Mean concentrations of ferrihydrite-iron in the deep gray matter reached over 500 µg/g wet weight and were three times higher compared to the temporal cortex, which already exceeded the healthy situation by six times. The amount of Fe^3+^ ions, detectable by EPR, and maghemite-iron was 1.7 times more abundant in the temporal cortex of the patient with aceruloplasminemia compared to the control subjects. Although the ferrihydrite/maghemite-iron ratio was highly variable among structures, ferrihydrite-iron was at least three orders of magnitude more abundant than maghemite-iron, on average.

The distribution of Fe^3+^ ions and iron mineralization products in the aceruloplasminemia brain is visualized in Figure 5. The globus pallidus and medial division of the thalamus, together with regions in the brain stem and cerebellum, showed the highest EPR-detectable Fe^3+^ concentration. The distribution of maghemite-iron appeared partially in accordance with this pattern, with the medial division of the thalamus, red nucleus, substantia nigra and dentate nucleus being the richest in this mineral form of iron. Maghemite levels in the lateral division of the thalamus and the putamen were below the detection limit of the technique.

**Figure 5.**
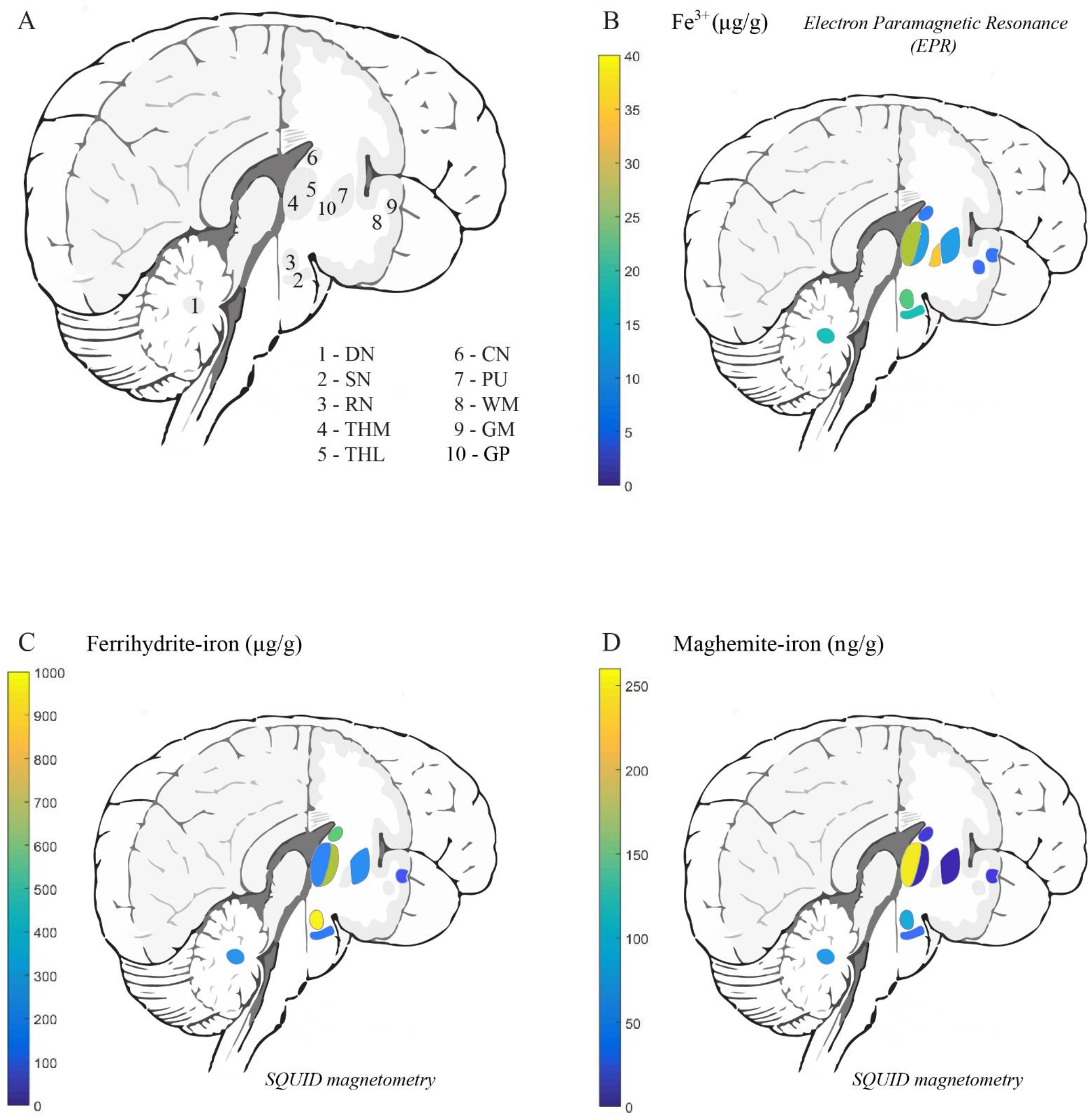
Distribution of different iron forms in aceruloplasminemia brain tissue. Abbreviations: DN – dentate nucleus, SN – substantia nigra, RN – red nucleus, THM – medial division of the thalamus, THL – lateral division of the thalamus, CN – caudate nucleus, GP – globus pallidus, PU – putamen, WM – white matter, GM – gray matter.

## Discussion

In this work, the iron-sensitive metrics R_2_* and χ, obtained from 7 T MRI, were combined with Electron Paramagnetic Resonance (EPR) and SQUID magnetometry to specify and quantify the different iron forms in the aceruloplasminemia brain. We detected and quantified ferrihydrite-iron – which is a measure of iron stores in the brain, maghemite-iron – an iron form that is recently attracting the attention of MR-physicists and biologists due to its potential neurotoxicity and the influence that it may have on MRI signal dephasing, and an EPR-sensitive pool of Fe^3+^ ions.

The comparison between R_2_* and QSM maps indicated that a (para)magnetic source is driving the increase in relaxation rate in both gray and white matter structures in aceruloplasminemia. We ascribe this source to iron, mainly ferrihydrite-iron. The R_2_* values showed that the red nucleus, the lateral division of the thalamus, and the substantia nigra were heavily iron-loaded, which is in agreement with a recent *in vivo* study in aceruloplasminemia (17). From the QSM maps, an increase in the paramagnetic signal was only observable in a few selected areas. Although QSM has the potential to more specifically identify the underlying source of R_2_* increase, our reconstructions suffered from artefacts and the quality was not sufficient for quantitative analysis. These artefacts were likely caused by the large heterogeneity of iron accumulation within structures and the extremely short proton relaxation time as a result of the extremely high iron concentrations that impaired the detection of the signal. QSM reconstructions in aceruloplasminemia might therefore benefit from lower magnetic field strengths, to partially mitigate these effects.

Of the three iron forms detected in the aceruloplasminemia brain, more than 90% was found to be associated with the ferritin storage protein and hemosiderin (ferrihydrite-iron). These findings agree with earlier histochemical studies of aceruloplasminemia brain tissue that showed the abundance of iron aggregates rich in Fe^3+^ that strongly reacted to ferritin light chain antibody (8, 9), and observations in healthy brain tissue (33). The concentrations of ferrihydrite-iron in the basal ganglia of our patient with aceruloplasminemia, could not be appropriately compared with results that had been previously obtained from the basal ganglia of healthy subjects and patients with neuroferritinopathy (29), due to substantial methodological differences between studies. The amount of ferrihydrite-iron in the temporal cortex of aceruloplasminemia, though, was six times higher than in healthy subjects and almost four times higher compared to patients with advanced Alzheimer’s disease that were measured under identical conditions (18, 34). Although these comparisons are limited to the temporal cortex, which is on average three times less rich in iron than the deep gray matter structures in aceruloplasminemia, they illustrate once more the severity of iron overload in aceruloplasminemia.

The distribution of ferrihydrite-iron throughout the brain in aceruloplasminemia did not reflect the total iron distribution in healthy brain tissue. While in normal ageing ferrihydrite-iron is most abundant in the globus pallidus and substantia nigra (11), in this aceruloplasminemia brain very high levels of ferrihydrite-iron were detected in the red nucleus, caudate nucleus and thalamus. In addition, the thalamus showed a striking lateralization, with the lateral part being far more abundant in ferrihydrite-iron than the medial part of the structure. This may be explained by specific pathways that have been recently highlighted as possibly involved in iron transport between the caudate nucleus and the pulvinar nucleus within the lateral division of the thalamus (17). The internal medullary lamina of the thalamus might additionally act as a barrier to prevent penetration of iron into the medial part of the structure (35). Although it should be noted that the globus pallidus was not assessed by magnetometry in this study, focal demyelination and iron accumulation with a preference for the pulvinar thalamus and caudate nucleus has also been described in patients with multiple sclerosis (36, 37), in which neurodegeneration and atrophy of these structures has been associated with disease progression and cognitive disturbances (37). Although this suggests a specific role of the caudate nucleus and (pulvinar) thalamus in neuroinflammation and neurodegeneration, different patterns of iron accumulation have been observed in Parkinson’s disease, multiple system atrophy, progressive supranuclear palsy (38), and other NBIA disorders (39), of which the clinical relevance remains to be elucidated.

The above-mentioned lateralization within the thalamus, which was observed with quantitative MRI, EPR and SQUID magnetometry, is in accordance with previous *in vivo* MRI findings in aceruloplasminemia (2, 20, 35). The relatively high R_2_* values obtained from the lateral division of the thalamus appeared to be driven by the large amount of ferrihydrite-iron in this region, while the medial part of the thalamus showed a lower R_2_* value despite being richer in maghemite-iron and EPR-detectable Fe^3+^ ions. Although the magnetization of maghemite is over two orders of magnitude higher compared to the one of ferrihydrite, its influence on iron-sensitive MRI remained limited due to the much lower concentration of maghemite nanocrystals compared to ferrihydrite (40). Even though these observations do not exclude that ferrihydrite-iron and maghemite-iron may contribute together to the increases in tissue magnetic susceptibility, ferrihydrite-iron likely remains the main driver of MRI susceptibility contrast in extremely iron-loaded cases.

The potential role of maghemite-iron in the pathogenesis of aceruloplasminemia should not be discarded purely based on its limited contribution to the total brain iron pool. Absolute SIRM values detected at 100 K in aceruloplasminemia were approximately two orders of magnitude higher than those reported in healthy subjects, at room temperature (41), which suggests that also maghemite-iron levels may have increased as a result of iron overload in aceruloplasminemia. In addition, the concentration of maghemite-iron detected in the temporal cortex of aceruloplasminemia was almost twice that of healthy controls and advanced cases of Alzheimer’s disease that were measured under identical conditions (18, 34). In contrast to ferrihydrite-iron, the regional distribution of maghemite-iron in the aceruloplasminemia brain was not substantially different from that of healthy controls (41), with the cerebellar and brain stem nuclei being relatively rich in this magnetic carrier, although thalamic structures were not specifically investigated by previous studies (41). The preserved distribution of maghemite-iron throughout the severely diseased aceruloplasminemia brain rather questions its biological relevance, which has already been a subject of debate in other iron-related neurodegenerative conditions (18, 27, 29, 30, 42).

The EPR-detected Fe^3+^ ions are complementary to the two above-mentioned iron forms, and may account for a part of the labile iron pool, which is implicated in the formation of reactive oxygen species, and for monomeric iron sites in the ferritin shell that are responsible for ferroxidase activity. Although EPR also has the potential to detect ferrihydrite-iron and maghemite-iron, the signal from these minerals is optimal at higher temperatures than used here (43), and would require at least ten times higher iron concentrations than found in this study to be detectable by EPR. Given its low abundance and relatively small magnetic moment, it is unlikely that Fe^3+^ ions significantly affect the MRI signal in aceruloplasminemia.

Our work has some limitations, the first being the long-term formalin fixation of the tissue. Since formalin fixation is known to reduce total tissue iron content and may have a detrimental effect on protein conformations (44-47), it is likely that the presented absolute concentrations of Fe^3+^ ions, ferrihydrite-iron and maghemite-iron in the aceruloplasminemia brain are underestimated compared to *in vivo*. Therefore, future studies should aim to minimize the duration of formalin fixation (34), and ideally include both aceruloplasminemia and control brain tissue to enable more in-depth analyses of the exact relationship between the different molecular forms of iron that are present in healthy and severely diseased states. On the other hand, the spatial distribution of the different iron forms, as presented in our work, is not significantly affected by prolonged formalin fixation (41). This is confirmed by the non-uniform distribution of iron within the thalamus in our study, which is in accordance with previous *in vivo* MRI findings in aceruloplasminemia (2, 20, 35). In addition, the reported ferrihydrite-iron and maghemite-iron concentrations should be considered as approximate rather than absolute values, given the assumptions that had to be made to obtain these iron concentrations from the SIRM values. However, we believe that reporting iron concentrations instead of SIRM values can help guiding the interpretation of the results in the context of biology. Finally, since our results are based on a single patient study, generalization to other cases should be done carefully. It should be noted, though, that the rarity of aceruloplasminemia makes the tissue availability a challenge, as illustrated by the fact that only six other autopsy cases have been published over decades (3, 8, 9, 48).

In conclusion, ferrihydrite-iron is the major determinant of iron-sensitive MRI contrast, and is the most abundant iron form in the aceruloplasminemia brain. Among the studied regions, the lateral division of the thalamus, the red nucleus and the basal ganglia contained the highest amounts of ferrihydrite-iron. The maintenance of ferritin as the major iron storing protein in extremely iron-loaded brain tissue, as observed in our study, has major implications for the concept of iron chelation as the main treatment for aceruloplasminemia and other NBIA disorders. In particular, deferiprone is currently being exploited to treat several NBIA diseases (22, 49). Deferiprone is the only iron chelator known to date that is capable of transporting iron across cell membranes and across the blood brain barrier. As it selectively binds Fe^3+^ and is capable of directly removing Fe^3+^ ions from the core of ferritin (50), our findings pathophysiologically support the clinical benefit of deferiprone that has been reported in patients with aceruloplasminemia and other NBIA disorders (22, 49).

## Data Availability

Data available upon request of the corresponding author

## Author contributions

**Lena H.P. Vroegindeweij:** Conceptualization, Methodology, Resources, Writing – Original Draft, Visualization, Project Administration; **Lucia Bossoni:** Methodology, Software, Investigation, Writing – Original Draft, Visualization, Project Administration; **Agnita J.W. Boon:** Conceptualization, Writing – Review & Editing; **J.H. Paul Wilson:** Conceptualization, Writing – Review & Editing; **Marjolein Bulk:** Resources, Writing – Review & Editing; **Martina Huber:** Resources, Writing – Review & Editing; **Jacqueline Labra-Muñoz:** Resources, Investigation, Writing – Review & Editing; **Andrew Webb:** Resources, Writing – Review & Editing; **Louise van der Weerd:**Methodology, Writing – Review & Editing, Supervision; **Janneke G. Langendonk:** Conceptualization, Methodology, Writing – Review & Editing, Supervision.

## Acknowledgements

The authors are very grateful to the patient with aceruloplasminemia and his family to agree with this research, would like to thank A. Lefering for access to the SQUID and technical assistance, and S.A. Bonnet and S. Zheng for access to the ICP-MS and measuring the iron concentration of the formalin solution.

