## Supplementary Materials for "Quantification of different iron forms in the aceruloplasminemia brain to explore iron-related neurodegeneration"

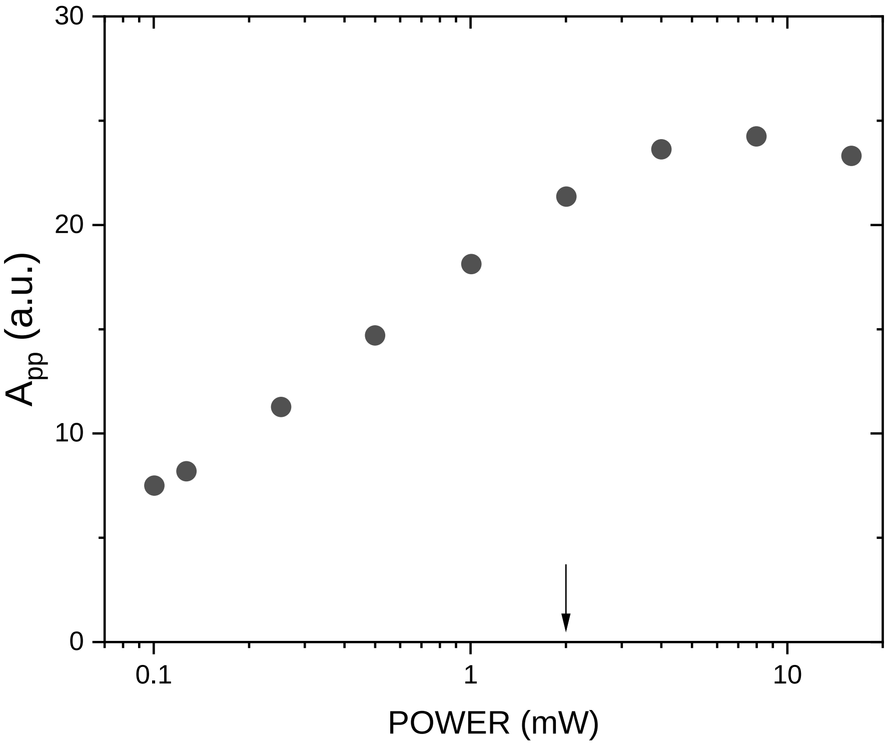


***Figure S1 Power attenuation experiment on the g'=4.3 band in one of the samples.*** *The amplitude peak-to-peak of the g'=4.3 band is shown as a function of the microwave power. The solid arrow shows the working point of the experiments presented in this work. Spectra were acquired at 6 K.*

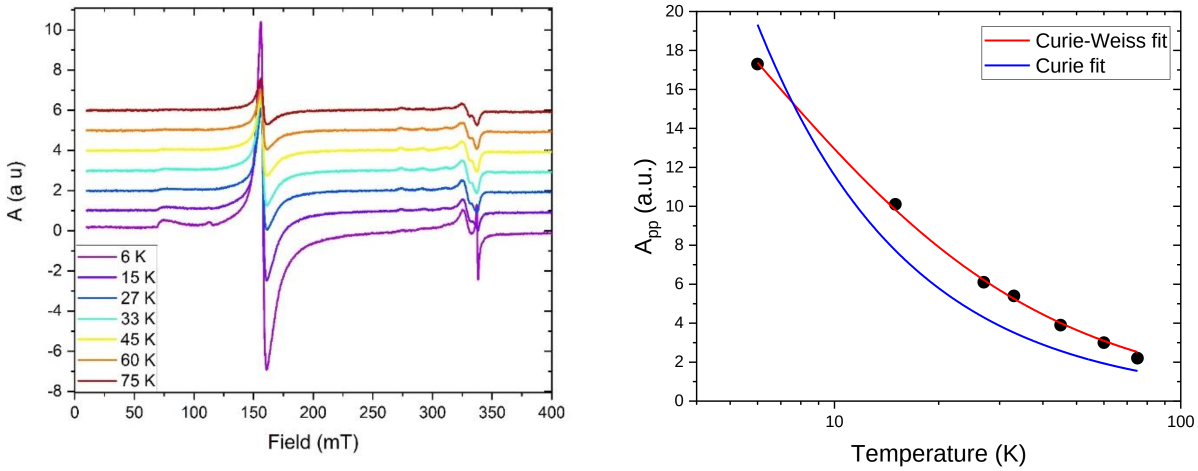


***Figure S2. Results of the temperature-dependent study of the putamen tissue block.*** *The left panel shows the spectra acquired at different temperatures in identical experimental conditions. The right panel shows the amplitude peak-to-peak of the g'=4.3 band (black circles) and the fit of the data with the Curie law (blue line) and the Curie-Weiss law (red line). The Curie-Weiss temperature extracted from the fit was 5.7 ±0.4 K.*

### Fit of the EPR spectra

Baseline-corrected EPR spectra were fit in EasySpin to the Hamiltonian below

$$\hat{H} = g \cdot \mu_B (B \cdot S) + D\left(S_z^2 - S(S+1)/3\right) + E\left(S_x^2 - S_y^2\right)$$

where g is the Landé factor, $\mu_B$ the Bohr magneton, **B** is the applied field and **S** the spin operator. The two final terms represent the zero-field splitting, where D is the axial splitting, and E the rhombic splitting.

An example of data fitting and best fitting parameters is reported below. The second integral of the fitted spectrum was used to estimate iron concentrations in this work.

***Table S1.***
***Best fitting parameters of the high-spin Fe(III) Hamiltonian $\hat{H}$, for the medial division of the thalamus.***

| $g_x$, $g_y$, $g_z$ | D (MHz) | E/D | $g_x$-strain, $g_y$-strain, $g_z$-strain |
|---|---|---|---|
| 1.80453, 1.98151, 2.01022 | 20960 | 0.3324 | 1.14746, 0.0961731, 0.0135024 |

*The large g-strain reflects the large distribution of Fe(III)-binding sites.*

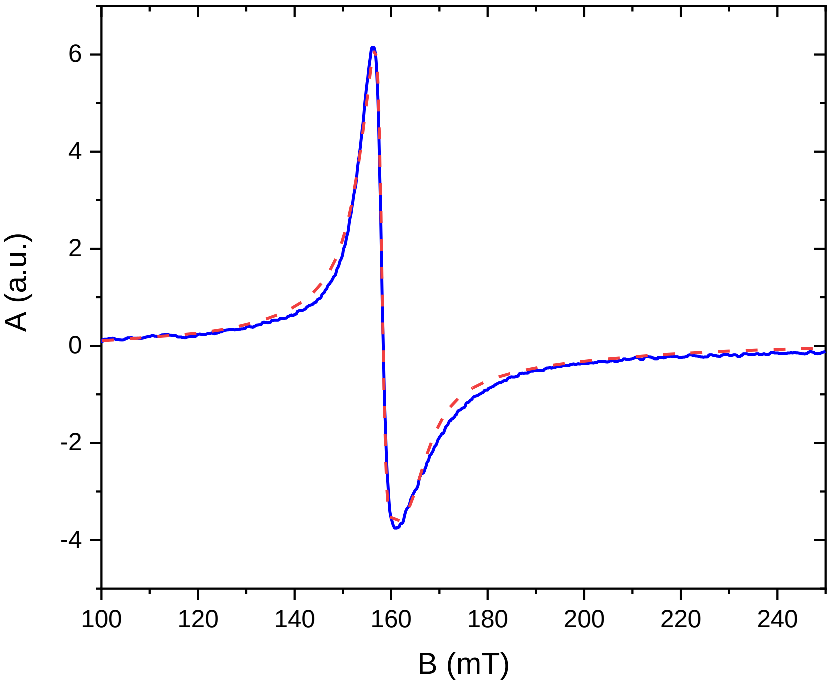


***Figure S3. EPR spectrum acquired on the medial division of the thalamus at 6 K (blue line).*** *Fit to the high-spin rhombic iron in red. The best fitting parameters are reported in Table1.*
